# Preterm birth increases placental expression of multidrug resistance (MDR) transporters irrespective of prepregnancy body mass index (BMI)

**DOI:** 10.1101/2021.07.30.21261204

**Authors:** Hailey Scott, Lilian M Martinelli, Enrrico Bloise, Kristin L Connor

## Abstract

**Context:** Preterm birth (PTB) and suboptimal prepregnancy body mass index (BMI) operate through inflammatory pathways to impair fetoplacental development. Placental efflux transporters mediate fetal protection and nutrition, however few studies consider the effect of both PTB and BMI on fetal protection. We hypothesized that PTB would alter the expression of placental multidrug resistance (MDR) transporters and selected pro-inflammatory cytokines, and that maternal underweight and obesity would further impair placental phenotype.

**Objective:** To determine whether placental MDR transporters P-glycoprotein (P-gp, encoded by *ABCB1*) and breast cancer resistance protein (BCRP/*ABCG2*), and pro-inflammatory cytokine levels are altered by PTB and maternal BMI.

**Design and Outcomes:** A cross-sectional study was conducted to assess the effect of PTB (+/- chorioamnionitis), or the effect of maternal prepregnancy BMI on placental MDR transporter and interleukin [IL]-6 and 8 expression in 60 preterm and 36 term pregnancies.

**Results:** *ABCB1* expression was increased in preterm compared to term placentae (p=0.04). P-gp (p=0.008) and BCRP (p=0.01) immunolabeling was increased among all preterm compared to term placentae, with P-gp expression further increased in preterm pregnancies with chorioamnionitis (PTC, p=0.007). Placental *IL-6* mRNA expression was decreased in PTC compared to term placentae (p=0.0005), and PTC was associated with the greatest proportion of anti-inflammatory medications administered during pregnancy. Maternal BMI group did not influence placental outcomes.

**Conclusions:** PTB and infection, but not prepregnancy BMI, alter placental expression of MDR transporters and *IL-6*. This may have implications for fetal exposure to xenobiotics that may be present in the maternal circulation in pregnancies complicated by PTB.

## Introduction

Preterm birth (PTB) is a leading cause of perinatal morbidity/mortality and death among children under five years globally^1^. Infants born preterm are at increased risk for both immediate and longer term health complications, including neurodevelopmental and sensory disabilities^2, 3^, mood disorders, as well as neurological^3, 4^ and cardiometabolic diseases later in life^5, 6^. PTB and adverse offspring development have been associated with inflammation and infection^7^, however, the etiology of PTB is not well understood.

Maternal underweight and obesity are also prevalent obstetric conditions^8-10^, and can inflict adverse effects on placental and fetal development, such as maternal vascular malperfusion, placental insufficiency, fetal growth restriction, and neurodevelopmental impairment^11-15^. These conditions are associated with chronic, low-grade inflammation^13^, and suboptimal quality and quantity of nutrients available to support pregnancy nutritional demands^14^. Despite the increasing burden of these common conditions, little is known about the mechanisms that contribute to poor fetoplacental development and adverse pregnancy outcomes such as PTB in pregnancies complicated by maternal malnutrition or suboptimal metabolic status.

Both PTB and suboptimal maternal body mass index (BMI) may operate through inflammatory pathways to impair fetoplacental development. The placenta is a critical regulator of the fetal environment, adjusting nutrient availability to the fetus while simultaneously protecting against the entry of microbes, xenobiotics and toxins throughout pregnancy^4, 16, 17^. Inflammation may impair placental function, including its ability to regulate the efflux of potentially harmful substances out of the placenta, or to undergo adaptations to protect the developing fetus from environmental insults^11^. Early activation of inflammatory pathways, including an increase in pro-inflammatory mediators in the placenta such as interleukin [IL]-6 and IL-8, may also lead to uterine contractions, cervical ripening, and membrane rupture^18^, culminating in PTB^7^. Further, preterm pregnancies with chorioamnionitis (inflammation of the fetal membranes) may be at risk for altered placental parameters such as placental transport, that may contribute to placental barrier dysregulation and intrauterine inflammation. Increased gestational tissue inflammation in chorioamnionitis may adversely affect offspring growth^19^ in addition to the developmental consequences associated with reduced gestation length alone. The inflammatory state associated with maternal underweight and obesity also favours increased inflammation of the placenta^14^, thus pregnancies complicated by suboptimal maternal BMI and PTB may be at greatest risk for adverse outcomes.

Placental efflux transporters regulate an array of potentially harmful endogenous and exogenous substances to mediate fetal protection and nutrition^20^. The function of these transporters is regulated in part by inflammatory molecules^17^. Commonly studied in the placenta are the multidrug resistance (MDR) transporters P-glycoprotein (P-gp) and breast cancer resistance protein (BCRP), which belong to the ATP-binding cassette (ABC) superfamily of efflux transporters^17^. P-gp is localized to the syncytiotrophoblast and cytotrophoblast cell layers of the placenta, and regulates the efflux of endogenous and synthetic glucocorticoids (cortisol; cortisone; prednisone), estrogens and progestogens, cytokines/chemokines (IL-6; interferon [IFN]-γ; tumor necrosis factor [TNF-α]), antibiotics (amoxicillin; doxorubicin; levofloxacin; rifampicin), and antiretrovirals^17, 21^. BCRP is localized to the syncytiotrophoblast, cytotrophoblast, and endothelial cells of fetal blood vessels, and transports endogenous steroids, antibiotics (beta-lactams; doxorubicin; fluoroquinolones), antivirals, sulfonylureas, nutrients, and toxins from the fetus back into the maternal circulation^17, 21^. The expression of placental P-gp and BCRP transporters varies throughout gestation, and may be higher in preterm placentae, and decrease with gestational age^22^. The effects of maternal BMI on the expression of placental efflux transporters are not well understood, but are critical to study, as in many regions of the world these conditions coexist with other infectious and inflammatory diseases and increased medication use, for which MDR transporters may be affected. Altered expression of these transporters with gestational age or suboptimal maternal BMI may impair the placenta’s ability to protect the developing fetus from harmful environmental conditions, potentially contributing to adverse pregnancy and infant outcomes.

Our primary study objective was to determine the effect of PTB on expression levels of MDR transporters and select inflammatory cytokines in the placenta. As both PTB and suboptimal maternal BMI are associated with increased inflammation and adverse pregnancy and infant outcomes, we then assessed the effects of maternal BMI on placental efflux transport potential within term and preterm pregnancies. We hypothesized that PTB would be associated with altered placental efflux transport and increased levels of pro-inflammatory cytokines, and that preterm pregnancies complicated by maternal underweight and obesity would show greatest changes in placental MDR expression levels.

## Methods

### Study population

Gestational tissue samples and clinical data were collected from 96 mother-infant dyads from the Research Centre for Women’s and Infants’ Health BioBank (RCWIH) at Mount Sinai Hospital, Toronto, Canada. Women delivered at term, preterm without chorioamnionitis, or preterm with chorioamnionitis, and were classified as underweight (UW), normal weight (NW), overweight (OW), or having obesity (OB) by prepregnancy BMI according to the World Health Organization and Society of Obstetricians and Gynaecologists of Canada^23^ guidelines, with the exception of women classified as underweight. Due to a low prevalence of women considered underweight in the study region, a prepregnancy BMI of less than 19 was considered to be underweight, 19 to 24.9 normal weight, 25.0 to 29.9 overweight, and 30.0 or greater obese. Among 60 preterm pregnancies, 12 women were classified as underweight by prepregnancy BMI, 17 as normal weight, 18 as overweight, and 13 as having obesity, and among 36 term pregnancies, 9 were classified as underweight, 7 normal weight, 9 overweight, and 11 obese. Based on current recommendations from the Institute of Medicine (IOM) guidelines (2009), maternal weight gain was categorized as inadequate, adequate, or excessive for singleton pregnancies based on prepregnancy BMI, where the recommended weight gain ranges are 12.5-18 kilograms, 11.5-16 kilograms, 7-11.5 kilograms or 5-9 kilograms^24^ for mothers who are underweight, normal weight, overweight or who have obesity, respectively. Study inclusion criteria included singleton pregnancies and live birth with no known fetal anomalies. As our aim is to understand how PTB and low and high maternal BMI influence pregnancy outcomes and program development, our cohort exclusion criteria comprised comorbidities that may potentially confound data interpretation due to their known effects on placental development and function and the inflammatory milieu: gestational diabetes mellitus, hypertension (including pregnancy induced hypertension), lupus, antiphospholipid antibody syndrome, Crohn’s disease, ulcerative colitis, colitis, Guillain-Barré syndrome, sexually transmitted infections, gastritis, urinary tract infections, smokers, documented recreational drug use during pregnancy, HELLP syndrome, pelvic inflammatory disease, and *in vitro* fertilization. Placentae were collected and snap frozen or fixed for downstream molecular and histologic analyses, according to RCWIH placental collection and storage protocols. Clinical data from the pregnancies (e.g. maternal prepregnancy BMI, medication history, mode of delivery) were obtained from chart reviews.

### RNA extraction and qPCR

Total RNA was extracted from placental tissue using the QIAGEN RNeasy Mini Kit (CAT #74134) according to the manufacturer’s instructions. Placental RNA samples were quantified and assessed for purity by Nanodrop (DeNovix DS-11 Spectrophotometer). cDNA was synthesized from RNA samples using the Bio-Rad iScript gDNA Clear cDNA Synthesis Kit (CAT #1725035) according to the manufacturer’s instructions.

Real-time quantitative PCR (qPCR) was used to measure the relative placental mRNA expression levels of MDR transporter genes *ABCB1* (encoding P-gp) and *ABCG2* (encoding BCRP), and *IL-6* and *IL-8*, key pro-inflammatory cytokines involved in the pathogenesis of PTB^25-27^ and correlated to alterations in MDR (P-gp and BCRP) expression and/or function^28, 29^. Genes of interest were normalized to the geometric mean of three stably expressed reference genes, β-actin (*ACTB)*, glyceraldehyde 3-phosphate dehydrogenase *(GAPDH)*, and TATA-Box binding protein *(TBP)*. Primer sequences for the genes of interest and reference genes can be found in Supplementary Table 1. qPCR reactions were performed for each gene using 2 µl of cDNA in 8 µl of Master Mix, prepared with Bio-Rad SsoAdvanced Universal SYBR Green (CAT #1725274) and 5 µM forward and reverse primer mix. Standards, samples, and a non-template control were run in triplicate using a Bio-Rad CFX384 real-time PCR detection system with the following cycling conditions: 95°C for 30 seconds, 40 cycles of 95°C for five seconds and 60°C for 20 seconds, followed by a melt curve from 65°C to 95°C at 0.5°C increments for five seconds. Relative mRNA expression was calculated using the ΔΔCq method (Bio-Rad CFX Maestro 1.1 Version: 4.1.2433.1219).

### Immunohistochemistry

5 μm placental sections were dewaxed and rehydrated, followed by boiling/cooling series of antigen retrieval in 10 mM citric acid/sodium citrate solution at pH 6.0, by microwaving and cooling for 10 minutes on ice. Sections were washed twice with 1X PBS, then quenched in 0.3% hydrogen peroxide in 97% methanol for 30 minutes at room temperature (RT). Following two more washes in 1X PBS with 0.1% Tween, sections were blocked with serum-free protein block (DAKO) for one hour (RT). Placentae were then incubated with P-gp antibody (Abcam) at 1:400 dilution or BCRP antibody (Abcam) at 1:1200 dilution in antibody diluent (DAKO) at 4°C overnight. Antibody diluent was incubated with negative control sections instead of primary antibodies. Sections were washed three times in 1X PBS with 0.1% Tween, then incubated with Biotin-conjugated secondary antibody (SPD-060-Spring Bioscience) for one hour (RT). Following three more washes in 1X PBS with 0.1% Tween, streptavidin-horseradish peroxidase (SPD-060-Spring Bioscience) was then applied for one hour (RT). Sections were washed twice with in 1X PBS with 0.1% Tween, and DAB peroxidase substrate (Vector Laboratories) was applied for one minute (RT). Slides were counter-stained with haematoxylin (Sigma-Aldrich) prior to mounting.

Semi-quantitative scoring of staining immunolabeled area and intensity were performed for all samples across the syncytiotrophoblast, cytotrophoblast, endothelium, and connective tissue as previously described with adaptations^29^. Staining in immunolabeled areas was scored as absent (0), 1-25% (1), 26-50% (2), 51-75% (3) and 76-100% (4).

### Univariate analyses

Univariate analyses were conducted to evaluate differences in MDR transporter and inflammatory cytokine expression across gestational age and maternal BMI groups. Differences in MDR transporter and inflammatory cytokine expression levels were first assessed between term and preterm pregnancies (inclusive of infection), as expression levels of MDR transporters and inflammatory markers are known to change over the course of pregnancy^17, 30, 31^, and between term, preterm without chorioamnionitis, and preterm with chorioamnionitis pregnancies, to determine the effect of the additional impact of infection on these placental parameters. Next, the effect of maternal prepregnancy BMI, specifically maternal underweight, overweight, and obesity prepregnancy compared to normal weight as the reference category, on placental MDR transporter and cytokine expression levels was assessed in term and preterm pregnancies separately. Finally, placental data were stratified by fetal sex to assess sex differences in placental transport and inflammation in term and preterm pregnancies. Outcome measures were tested for normality and unequal variances (Levene test). Where possible, non-normal molecular data were log transformed to achieve normality. Differences between gestational age or maternal prepregnancy BMI groups and continuous outcome measures were determined by ANOVA with Tukey *post hoc* or Welch with Games-Howell *post hoc* for normally distributed data, or Kruskal–Wallis test with Steel–Dwass *post hoc* for nonparametric data. For categorical data, Likelihood Ratio Chi Square tests were used to evaluate the associations between gestational age or maternal BMI groups and categorical outcome variables, testing for differences in the distribution of participants by variable category across gestational age or maternal prepregnancy BMI group. Statistical significance was defined as p<0.05. Data were analysed using JMP statistical software (Version:14.0.0). Data are presented as median (interquartile range) for non-parametric continuous data, and frequency (percentage) for categorical variables.

### Multivariable analyses

We performed multivariable regression to determine the relationships between gestational age at delivery or maternal prepregnancy BMI with MDR transporter and inflammatory cytokine expression levels. Linear (Standard Least Squares) regression models were used to determine the associations between gestational age (continuous), or maternal BMI (continuous) and MDR transporter and inflammatory cytokine expression levels. For all BMI analyses, regressions were stratified by term and preterm birth. Data are reported as β (95% confidence interval) and p value from Standard Least Squares regression models. Covariates of interest were identified *a priori* and included gestational age (weeks), maternal prepregnancy BMI (continuous), infant sex, z-gestational weight gain, antibiotic use (yes/no), and chorioamnionitis status. An unadjusted model was first used to identify the associations between gestational age at delivery or prepregnancy BMI alone (as continuous variables) with placental MDR transporter and inflammatory cytokine expression levels. An adjusted model was then used to determine associations between gestational age at delivery or prepregnancy BMI with placental transporter and cytokine expression levels, adjusting for gestational age at delivery (weeks), maternal prepregnancy BMI (continuous), infant sex, z-gestational weight gain, antibiotic use, and chorioamnionitis status.

## Results

### Maternal clinical characteristics

Inclusive of BMI, mothers who delivered preterm were younger than mothers who delivered at term (preterm, PT: 32 years [29.3, 35]; term, T: 34 years [31.3, 37.8], p=0.005), and had lower present weight (PT: 73 kg [63.9, 84.5]; T: 84 kg [67.2, 95], p=0.02) and total gestational weight gain (PT: 9.3 kg [6.68, 12.3]; T: 14 kg [12, 18], p<0.0001) than mothers who delivered at term. There were no differences in prepregnancy weight or prepregnancy BMI between preterm and term pregnancies. More mothers with preterm births received antibiotics (PT: n=52, 86.7%; T: n=21, 58.3%, p=0.0002), glucocorticoids (PT: n=53, 88.3%; T: n=1, 2.78%, p<0.0001), and tocolytics (PT: n=22, 36.7%; T: n=0, p<0.0001) than term births. All mothers who delivered preterm and had chorioamnionitis received glucocorticoids, tocolytics, and/or antibiotics during pregnancy, followed by 97% of mothers who delivered preterm but did not have chorioamnionitis, and 58% of mothers who delivered at term (p<0.0001, Table 1). Specifically, 93% of mothers who delivered preterm and had chorioamnionitis were administered antibiotics, followed by 81% of preterm pregnancies without chorioamnionitis, and 58% of term pregnancies (p=0.0003, Table 1). No mothers who delivered at term received tocolytics. All (100%) preterm pregnancies with chorioamnionitis, 77% of preterm, and one pregnancy that delivered at term (3%) were administered glucocorticoids (p<0.0001, Table 1). Twenty-nine (48%) of preterm pregnancies recruited for this study had chorioamnionitis, and 31 (52%) were preterm pregnancies without chorioamnionitis. Of the preterm pregnancies with chorioamnionitis, fetal membrane inflammation grade (defined as 0 [no inflammation], 1 [moderate clusters of neutrophils], 2 [severe, confluent neutrophils between chorion and decidua], or 3 [microabscesses between chorion and decidua]) was mostly classified as inflammation grade 2 (47%), with one membrane classified as grade 3 (5.3%). Most preterm gestational tissues without infection (65%) and at term (89%) were classified as inflammation grade 0.

**Table 1.** Maternal antibiotics, tocolytics, and glucocorticoids administered during pregnancy among preterm pregnancies with chorioamnionitis, preterm pregnancies without chorioamnionitis, and term pregnancies.

|  | Preterm Chorio<br>(n=29) | Preterm<br>(n=31) | Term<br>(n=36) | p-value |
| --- | --- | --- | --- | --- |
| Any medication received<sup>1</sup> (n [%]) |  |  |  | **<0.0001** |
| Yes | 29 (100) | 30 (96.8) | 21 (58.3) |  |
| No | 0 | 1 (3.23) | 15 (41.7) |  |
| **Antibiotics** |  |  |  |  |
| Received antibiotics (n [%]) |  |  |  | **0.0003** |
| Yes | 27 (93.1) | 25 (80.6) | 21 (58.3) |  |
| No | 1 (3.45) | 4 (12.9) | 15 (41.7) |  |
| Missing | 1 (3.45) | 2 (6.45) | 0 |  |
| Antibiotics given<sup>2</sup> (n [%]) |  |  |  | - |
| Amoxicillin | 3 (11.1) | 1 (4.0) | 0 |  |
| Ampicillin | 7 (25.9) | 2 (8.0) | 0 |  |
| Ancef | 5 (18.5) | 2 (8.0) | 4 (19.0) |  |
| Cefazolin | 0 | 3 (12.0) | 7 (33.3) |  |
| Clindamycin | 3 (11.1) | 2 (8.0) | 0 |  |
| Erythromycin | 13 (48.1) | 9 (36.0) | 1 (4.76) |  |
| Flagyl | 1 (3.70) | 0 | 0 |  |
| Gentamycin | 6 (22.2) | 0 | 0 |  |
| Macrobid | 0 | 1 (4.0) | 0 |  |
| Penicillin G | 13 (48.1) | 10 (40.0) | 1 (4.76) |  |
| Tobramycin | 2 (7.41) | 0 | 0 |  |
| Vancomycin | 1 (3.70) | 2 (8.0) | 1 (4.76) |  |
| Antibiotic dose (mg)<sup>2</sup> |  |  |  | - |
| Amoxicillin | 1250 (875, 1625) | 2000 | - |  |
| Ampicillin | 2000 (1000, 2000) | 2000 | - |  |
| Ancef | 2000 (1625, 2000) | 2000 | 2000 |  |
| Cefazolin | - | 2000 (2000, 2500) | 2000 (2000, 2000) |  |
| Clindamycin | 900 | 900 | - |  |
| Erythromycin | 250 | 250 | . |  |
| Flagyl | - | - | - |  |
| Gentamycin | 80 | - | - |  |
| Macrobid | - | 5000 | - |  |
| Penicillin G<sup>3</sup> | 2500 (2500, 3750) | 5000 (2500, 5000) | . |  |
| Tobramycin | 80 | - | - |  |
| Vancomycin | 500 | 1000 | . |  |
| **Tocolytics** |  |  |  |  |
| Received tocolytics (n [%]) |  |  |  | **<0.0001** |
| Yes | 11 (37.9) | 11 (35.5) | 0 |  |
| No | 17 (58.6) | 20 (64.5) | 36 (100) |  |
| Missing | 1 (3.45) | 0 | 0 |  |
| Tocolytic type (n [%]) |  |  |  | 0.20 |
| MgSO<sub>4</sub> | 7 (63.6) | 4 (36.4) | - |  |
| MgSO<sub>4</sub> and Morphine | 1 (9.09) | 0 | - |  |
| Adalat | 3 (27.3) | 6 (54.6) | - |  |
| Adalat and Indomet | 0 | 1 (9.09) | - |  |
| **Glucocorticoids** |  |  |  |  |
| Received glucocorticoids (n [%]) |  |  |  | **<0.0001** |
| Yes | 29 (100) | 24 (77.4) | 1 (2.78) |  |
| No | 0 | 7 (22.6) | 35 (97.2) |  |
| Number of courses given | 2 (2, 2) | 2 (2, 2) | 2 | 0.67 |
| Missing | n=1 | n=2 | - |  |
| Gestational age given | 26 (24, 28)<sup>A</sup> | 31 (26, 32)<sup>B</sup> | 28<sup>AB</sup> | **0.03** |
| Missing | n=2 | n=1 | - |  |

Data are n (%) (Likelihood Ratio Chi Square test) for categorical variables or median (IQR; Kruskal-Wallis/Wilcoxon test for non-parametric data with Steel-Dwass *post-hoc*) for continuous variables. \*p<0.05. <sup>1</sup>Received maternal antibiotics, glucocorticoids, and/or tocolytics. <sup>2</sup>Some mothers were given multiple types of antibiotics. <sup>3</sup>Some mothers were given multiple doses of Penicillin G. Preterm Chorio = preterm with chorioamnionitis.

Among pregnancies that delivered preterm, most mothers who were underweight, normal weight, or had obesity prepregnancy had inadequate gestational weight gain, while most mothers classified as overweight prepregnancy had adequate weight gain based on IOM (2009) guidelines by prepregnancy BMI (p=0.02, Table 2). In contrast, few mothers at term had inadequate gestational weight gain; most mothers who were underweight prepregnancy were classified as having adequate weight gain, an equal number of mothers who were normal weight had adequate or excessive weight gain, and most mothers who were overweight or had obesity had excessive gestational weight gain (p=0.005, Table 3). Mothers with obesity who delivered preterm were also more likely to receive antibiotics during pregnancy: 100% of mothers with obesity received antibiotics, followed by 91.7% of mothers who were underweight, 88.2% who were normal weight, and 72.2% of mothers who were overweight (p=0.04, Table 2). There were no differences in antibiotic administration or other demographic or pregnancy characteristics among mothers who delivered at term by maternal BMI group (Table 3).

**Table 2.**
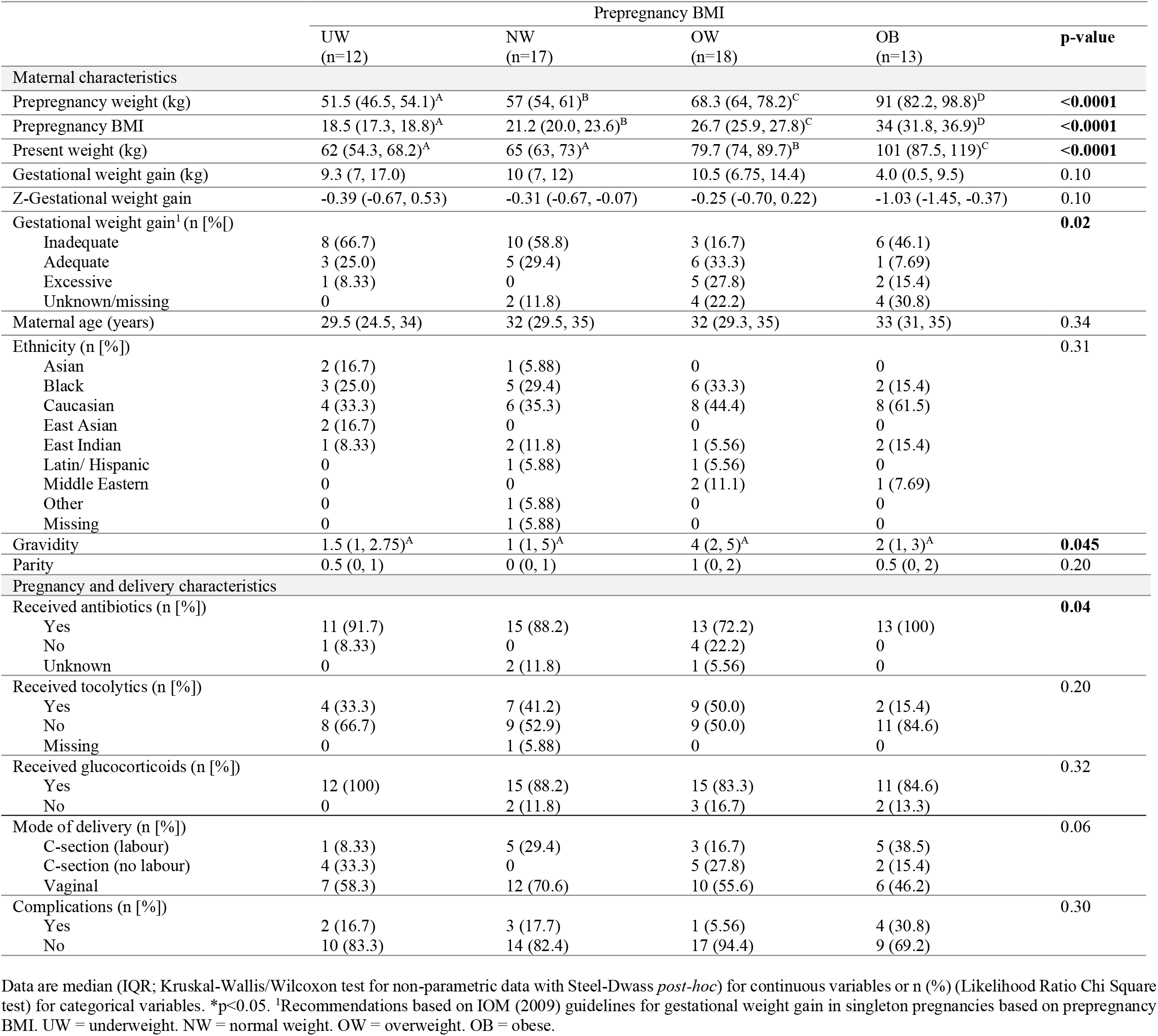
Maternal and pregnancy characteristics by prepregnancy BMI in preterm pregnancies.

**Table 3.** Maternal and pregnancy characteristics by prepregnancy BMI in term pregnancies.

|  | Prepregnancy BMI |  |  |  |  |
| --- | --- | --- | --- | --- | --- |
|  | UW<br>(n=9) | NW<br>(n=7) | OW<br>(n=9) | OB<br>(n=11) | p-value |
| Maternal characteristics |  |  |  |  |  |
| Prepregnancy weight (kg) | 50 (46.5, 54.3)<sup>A</sup> | 61 (59, 64)<sup>B</sup> | 71.4 (60.8, 79.8)<sup>B</sup> | 94 (76.5, 116)<sup>C</sup> | <0.0001 |
| Prepregnancy BMI | 18.4 (17.7, 18.7)<sup>A</sup> | 22.4 (21.1, 24.4)<sup>B</sup> | 26.3 (25.7, 27.5)<sup>C</sup> | 35.5 (31.7, 44.1)<sup>D</sup> | <0.0001 |
| Present weight (kg) | 64.5 (63.1, 66.7)<sup>A</sup> | 79 (72, 86)<sup>AB</sup> | 87.7 (74.4, 92.5)<sup>BC</sup> | 102 (90, 122)<sup>C</sup> | <0.0001 |
| Gestational weight gain (kg) | 14.6 ± 3.22 | 17.4 ± 5.98 | 17.4 ± 4.29 | 12.6 ± 5.32 | 0.13 |
| Z-Gestational weight gain | 0.24 ± 0.39 | 0.58 ± 0.72 | 0.57 ± 0.51 | 0.003 ± 0.64 | 0.13 |
| Gestational weight gain<sup>1</sup> (n [%]) |  |  |  |  | 0.005 |
| Inadequate | 1 (11.1) | 1 (14.3) | 0 | 0 |  |
| Adequate | 6 (66.7) | 3 (42.9) | 0 | 3 (27.3) |  |
| Excessive | 1 (11.1) | 3 (42.9) | 8 (88.9) | 7 (63.6) |  |
| Unknown/missing | 1 (11.1) | 0 | 1 (11.1) | 1 (9.09) |  |
| Maternal age (years) | 34.2 ± 3.70 | 33.4 ± 2.99 | 35.6 ± 3.40 | 34.2 ± 4.71 | 0.73 |
| Ethnicity (n [%]) |  |  |  |  | 0.20 |
| Asian | 4 (44.4) | 1 (14.3) | 2 (22.2) | 0 |  |
| Black | 0 | 0 | 2 (22.2) | 2 (18.2) |  |
| Caucasian | 3 (33.3) | 6 (85.7) | 4 (44.4) | 5 (45.5) |  |
| East Asian | 0 | 0 | 0 | 0 |  |
| East Indian | 1 (11.1) | 0 | 0 | 1 (9.09) |  |
| Latin/ Hispanic | 0 | 0 | 0 | 1 (9.09) |  |
| Middle Eastern | 0 | 0 | 0 | 1 (9.09) |  |
| Other | 0 | 0 | 0 | 1 (9.09) |  |
| Missing | 1 (11.1) | 0 | 1 (11.1) | 0 |  |
| Gravidity | 2 (1, 3) | 2 (1, 2) | 2 (1, 3.5) | 3 (2, 4) | 0.18 |
| Parity | 1 (0, 1) | 1 (0, 1) | 0 (0, 1) | 1 (1, 2) | 0.13 |
| Pregnancy and delivery characteristics |  |  |  |  |  |
| Received antibiotics (n [%]) |  |  |  |  | 0.24 |
| Yes | 4 (44.4) | 4 (57.1) | 4 (44.4) | 9 (81.8) |  |
| No | 5 (55.6) | 3 (42.9) | 5 (55.6) | 2 (18.2) |  |
| Unknown | 0 | 0 | 0 | 0 |  |
| Received glucocorticoids (n [%]) |  |  |  |  | 0.49 |
| Yes | 0 | 0 | 0 | 1 (9.09) |  |
| No | 9 (100) | 7 (100) | 9 (100) | 10 (90.1) |  |
| Mode of delivery (n [%]) |  |  |  |  | 0.12 |
| C-section (labour) | 0 | 0 | 1 (11.1) | 2 (18.2) |  |
| C-section (no labour) | 3 (33.3) | 4 (57.1) | 2 (22.2) | 7 (63.6) |  |
| Vaginal | 6 (66.7) | 3 (42.9) | 6 (66.7) | 2 (18.2) |  |
| Complications (n [%]) |  |  |  |  | 0.53 |
| Yes | 2 (22.2) | 3 (42.9) | 1 (11.1) | 3 (27.3) |  |
| No | 7 (77.8) | 4 (57.1) | 8 (88.9) | 8 (72.7) |  |

Data are means ± SD (ANOVA; normal distribution/equal variance) or median (IQR; Kruskal-Wallis/Wilcoxon test for non-parametric data with Steel-Dwass *post-hoc*) for continuous variables. Data are n (%) (Likelihood Ratio Chi Square test) for categorical variables. \*p<0.05. <sup>1</sup>Recommendations based on IOM (2009) guidelines for gestational weight gain in singleton pregnancies based on prepregnancy BMI. UW = underweight. NW = normal weight. OW = overweight. OB = obese.

### Placental IL-6 mRNA expression was decreased with preterm birth and chorioamnionitis infection

Placental *IL-6* mRNA expression levels were decreased in all preterm (with and without chorioamnionitis) compared to term pregnancies (p=0.0002, Figure 1A), and levels increased with advancing gestational age (unadjusted model: β=0.04 [0.02, 0.06], p=0.001). When stratifying by term and preterm +/- chorioamnionitis status, placental *IL-6* mRNA expression levels were lower in preterm pregnancies with chorioamnionitis compared to term pregnancies (p=0.0005, Figure 1A). There was no effect of gestational age or chorioamnionitis status on placental *IL-8* mRNA expression. Placental cytokine expression levels were also not different across maternal BMI groups at preterm or term (Supplementary Table 2), or when comparing male and female placentae at preterm or term (Supplementary Table 3).

**Figure 1.**
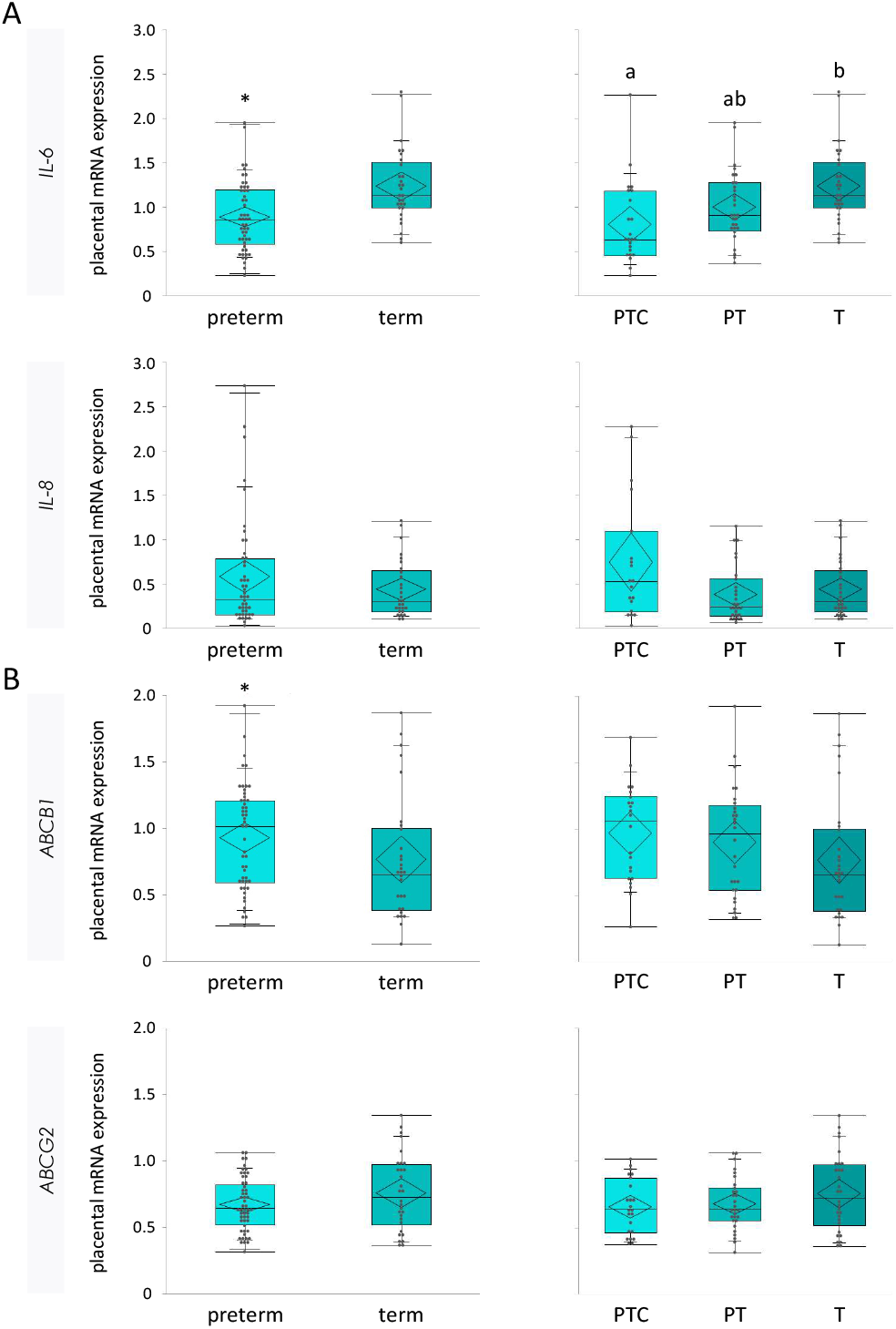
Placental mRNA expression levels of (A) *IL-6* and *IL-8* pro-inflammatory cytokines and (B) *ABCB1* and *ABCG2* efflux transporters. Placental *IL-6* mRNA expression levels were decreased in all preterm (with and without chorioamnionitis) compared to term placentae (preterm: 0.85 [0.58, 1.20]; term: 1.13 [0.99, 1.50], p=0.0002), and further decreased in preterm pregnancies with chorioamnionitis compared to term placentae (PTC: 0.63 [0.45, 1.18]^A^; PT: 0.90 [0.73, 1.27]^AB^; T: 1.13 [0.99, 1.50]^B^, p=0.0005). There was no effect of gestational age or chorioamnionitis status on placental *IL-8* mRNA expression. Placental *ABCB1* mRNA expression levels were increased in all preterm compared to term placentae (preterm: 1.02 [0.59, 1.21]; term: 0.65 [0.38, 1.00], p=0.04), but there was no effect of chorioamnionitis. Placental mRNA expression levels of *ABCG2* did not differ by gestational age or infection status. Data are median [IQR] with 95% confidence diamonds and p-value from ANOVA, or Welch test with Games-Howell post-hoc, *p<0.05. *IL-8* = interleukin 8. *IL-6* = interleukin-6. *ABCB1* = gene encoding P-glycoprotein. *ABCG2* = gene encoding breast cancer resistance protein. PTC = preterm with chorioamnionitis. PT = preterm without chorioamnionitis. T = term.

### MDR transporter mRNA and protein expression were increased in preterm placentae

Placental *ABCB1* mRNA expression levels were increased in all preterm pregnancies (with and without of chorioamnionitis) compared to term pregnancies (p=0.04, Figure 1B). mRNA expression levels of *ABCG2* did not differ by gestational age. There was also no effect of chorioamnionitis status on placental *ABCB1* or *ABCG2* mRNA expression levels (Figure 1B).

Placental P-gp (PT: score of 3 [1, 4]; T: score of 1 [0, 3.25], p=0.008) and BCRP (PT: score of 4 [4, 4]; T: score of 4 [4, 4], p=0.01) protein immunolabeled areas of the syncytiotrophoblast (but not other layers of the placenta) were also increased among all preterm pregnancies compared to term pregnancies. Further, there was an effect of infection status on P-gp protein, where syncytiotrophoblast immunolabeled area of P-gp was increased in preterm pregnancies with chorioamnionitis compared to term pregnancies (p=0.007, Figure 2A-B, Figure 3). There was also a small statistical difference in BCRP syncytiotrophoblast immunolabeled area across gestational age and infection groups, though the median staining value for all groups was 4 (76-100% of immunolabeled area staining): all placentae from preterm pregnancies without chorioamnionitis demonstrated 76-100% area stained, and thus scored with a 4, while staining in term pregnancies ranged from 0 (absent) to 4 (76-100% of area), suggesting increased BCRP staining in preterm placentae without chorioamnionitis (p=0.03, Figure 2A-B, Figure 4).

**Figure 2.**
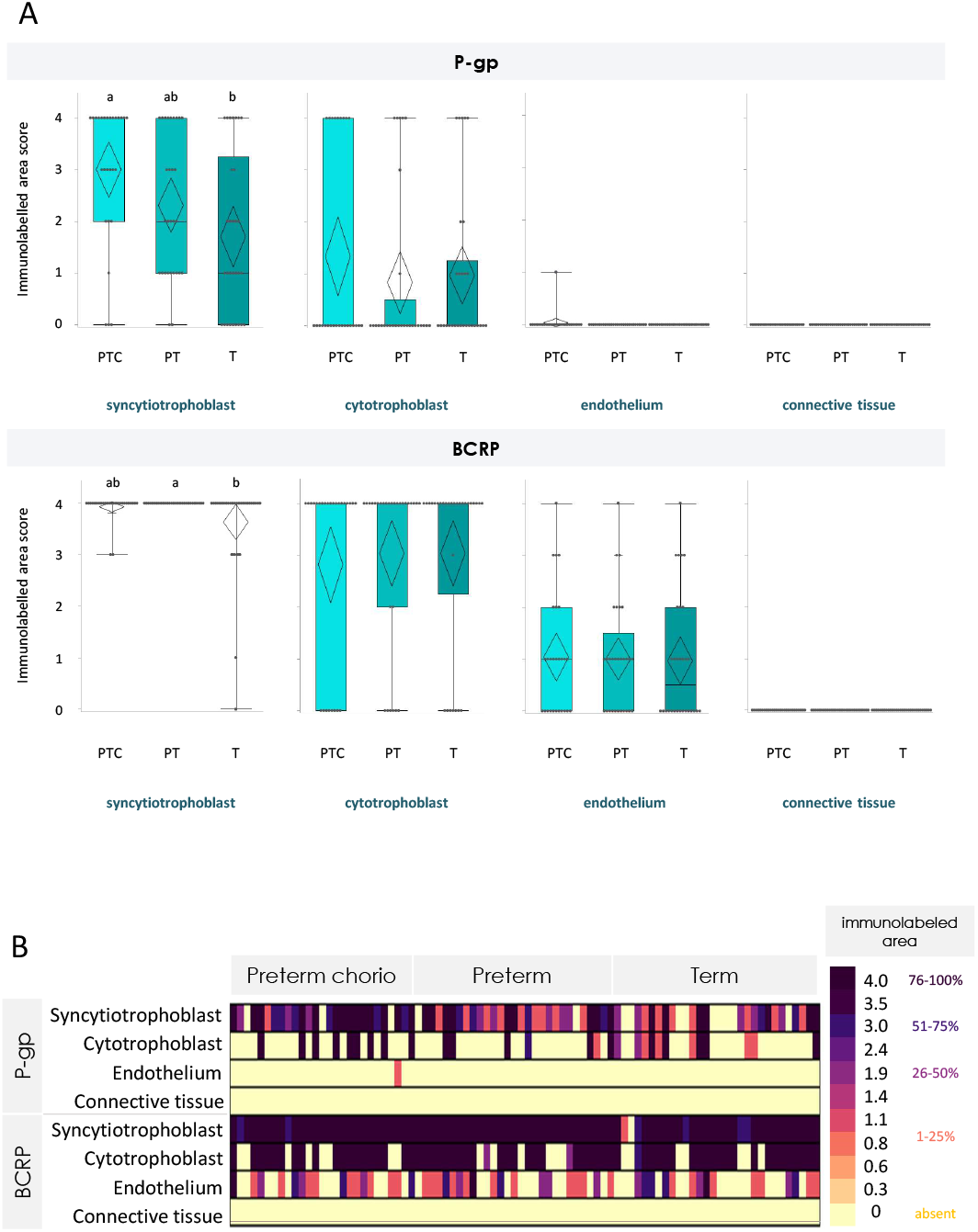
Placental P-gp and BCRP protein staining of immunolabeled areas in preterm pregnancies with chorioamnionitis, preterm pregnancies without chorioamnionitis, and term pregnancies, across four placental regions. (A) Box plots of placental P-gp and BCRP staining of immunolabeled areas in preterm pregnancies with chorioamnionitis (PTC), preterm pregnancies without chorioamnionitis (PT), and term pregnancies (T). P-gp immunolabeled area staining of the syncytiotrophoblast was increased in preterm pregnancies with chorioamnionitis compared to term pregnancies. BCRP immunolabeled area staining of the syncytiotrophoblast was increased in preterm pregnancies without chorioamnionitis compared to term pregnancies. There were no differences in P-gp or BCRP immunolabeling of the cytotrophoblast, fetal vessel endothelium, or connective tissue stratified by gestational age and infection group. Staining was scored as absent (0), 1-25% (1), 26-50% (2), 51-75% (3) and 76-100% (4). Semi-quantitative analysis of staining in immunolabeled area data were analysed using Kruskal–Wallis test with Steel–Dwass *post hoc*. p<0.05. (B) Heatmap of staining of immunolabeled areas in preterm pregnancies with chorioamnionitis (Preterm chorio), preterm without chorioamnionitis, and term pregnancies. P-gp = P-glycoprotein. BCRP = breast cancer resistance protein.

**Figure 3.**
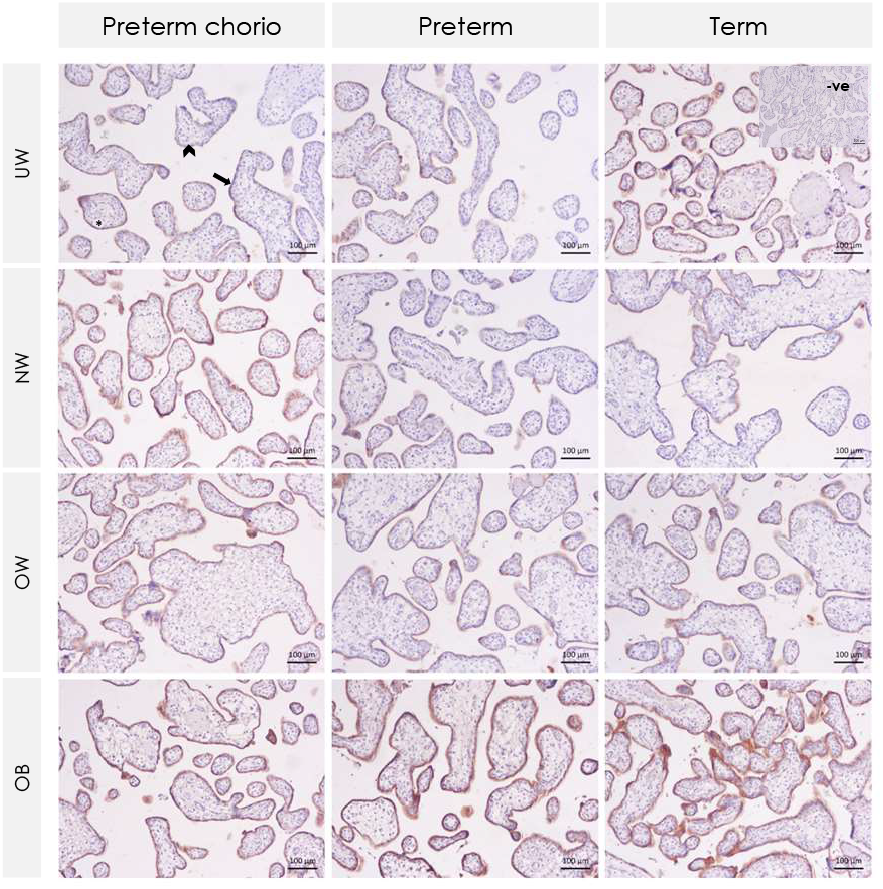
Representative IHC images of P-gp staining in human placentae across maternal prepregnancy BMI groups stratified by gestational age and infection status. Arrow indicates immunolabeled syncytiotrophoblast. Arrowhead indicates unlabeled cytotrophoblast. Asterisk indicates fetal blood vessels with no labeled endothelium. P-gp = P-glycoprotein. Preterm chorio = preterm with chorioamnionitis. UW = underweight. NW = normal weight. OW = overweight. OB = obese.

**Figure 4.**
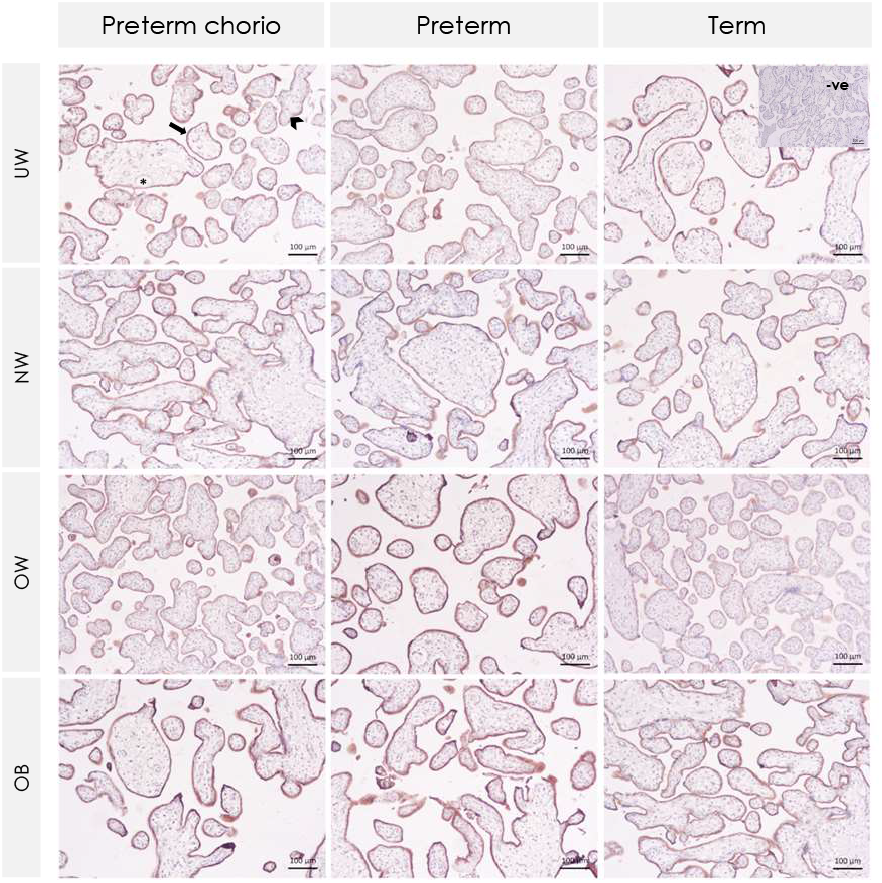
Representative IHC images of BCRP staining in human placentae across maternal prepregnancy BMI groups stratified by gestational age and infection status. Arrows indicate labeled syncytiotrophoblast. Arrowhead indicates cytotrophoblast. Asterisks indicate fetal blood vessels with no labeled endothelium. BCRP = Breast cancer resistance protein. Preterm chorio = preterm with chorioamnionitis. UW = underweight. NW = normal weight. OW = overweight. OB = obese.

There was no effect of maternal BMI group on placental mRNA or protein expression of *ABCB1*/P-gp or *ABCG2*/BCRP among term or preterm pregnancies (Supplementary Table 2, Figure 5A-C). When considering maternal BMI as a continuous variable, placental mRNA expression of *ABCG2*, but not *ABCB1*, increased with increasing maternal BMI at preterm (unadjusted and adjusted β=0.008 [0.0001, 0.02], p=0.047). There were no differences in placental *ABCG2* or *ABCB1* mRNA expression by maternal BMI (continuous) at term. There was no effect of placental sex on mRNA or protein expression of *ABCB1*/P-gp or *ABCG2*/BCRP among term or preterm pregnancies (Supplementary Table 3).

**Figure 5.**
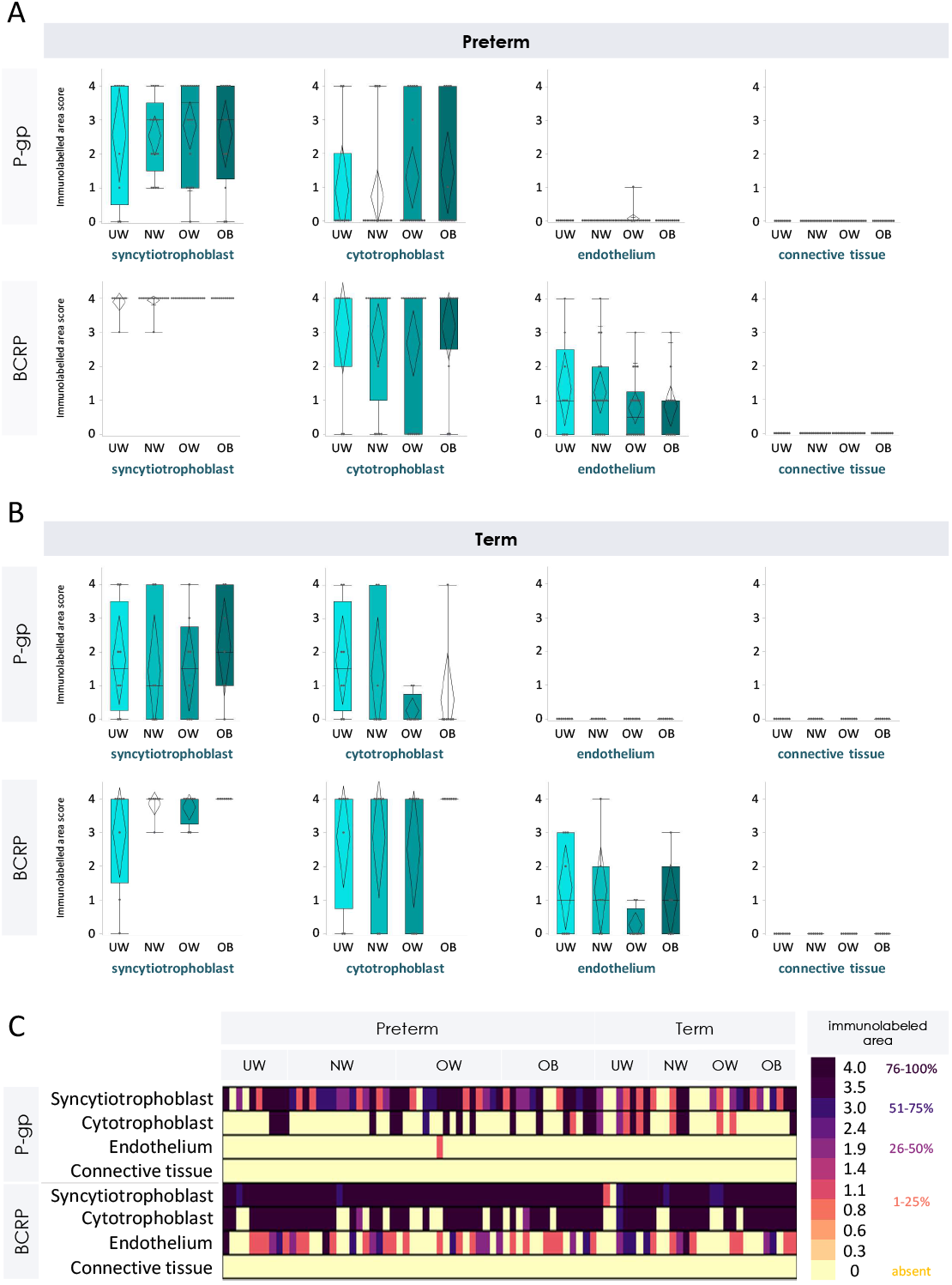
Placental P-gp and BCRP protein staining of immunolabeled area in pregnancies with maternal underweight, normal weight, overweight, or obesity prepregnancy stratified by preterm and term pregnancies, across four placental regions. Box plots of P-gp and BCRP staining of immunolabeled areas by maternal prepregnancy BMI group in preterm (A) and term (B) pregnancies. There were no differences in placental P-gp or BCRP immunolabeled area staining by maternal prepregnancy BMI group at preterm or term. Staining was scored as absent (0), 1-25% (1), 26-50% (2), 51-75% (3) and 76-100% (4). Semi-quantitative analysis of staining in immunolabeled area data were analysed using Kruskal–Wallis test. p<0.05. (C) Heatmap of staining of immunolabeled areas across maternal prepregnancy BMI group in preterm and term pregnancies. P-gp = P-glycoprotein. BCRP = Breast cancer resistance protein. Preterm chorio = preterm with chorioamnionitis. UW = underweight. NW = normal weight. OW = overweight. OB = obese.

## Discussion

We examined the effect of PTB and maternal prepregnancy BMI on placental expression levels of MDR transporters and key inflammatory cytokines to better understand the placental mechanisms through which these common conditions may lead to poor pregnancy outcomes. We found a gestational age effect, where placental *ABCB1*/P-gp mRNA and protein expression levels were increased, and placental *IL-6* mRNA expression levels were decreased, in all preterm pregnancies compared to term pregnancies, effects that were amplified in preterm pregnancies with chorioamnionitis. Mothers who delivered preterm with chorioamnionitis also received the greatest proportion of glucocorticoids, antibiotics, and tocolytics during pregnancy, which may explain reductions in placental *IL-6* mRNA expression levels. There was no effect of maternal BMI group or placental sex on expression levels of placental MDR transporters or select inflammatory cytokines among term or preterm pregnancies, however there was a small significant difference where placental mRNA expression of *ABCG2* increased with increasing maternal BMI at preterm. PTB and chorioamnionitis thus alter the expression of key placental transporters and inflammatory regulators, which may impact placental development and function.

Our observations that placental transporter expression levels were increased in all preterm pregnancies is consistent with previous findings that suggest the expression of P-gp and BCRP transporters in the placenta decrease with gestational age^22, 32^. Placental *ABCB1* mRNA and syncytiotrophoblast P-gp protein levels are higher in first trimester compared to term^30, 33^, supporting the important role of P-gp in fetal exchange and protection, especially at earlier stages of pregnancy^30^. In contrast, no differences in *ABCG2* mRNA levels were found across gestation. We did, however, find increased BCRP protein expression in preterm placentae, suggesting a disconnect between gene and protein level results, which could be in part explained by the semi-quantitative nature of immunohistochemistry, or may suggest a functional adaptation in protein translation in response to other regulating factors, such placental oxygen levels^30^. Placental P-gp and BCRP expression have also been shown to be increased in mothers exhibiting high white blood cell count, which may be an indication of infection^22^, in line with our findings of increased transporter expression in preterm pregnancies with chorioamnionitis. While results were not significant at the mRNA level, placental P-gp protein expression in the syncytiotrophoblast was highest in preterm pregnancies with chorioamnionitis compared to term pregnancies. In contrast, other studies have shown increased *ABCB1* and *ABCG2* mRNA and BCRP protein levels, but decreased P-gp protein levels with chorioamnionitis, which could be explained by post-transcriptional influences of inflammation^29^. Our results suggest that P-gp protein expression is not only increased in preterm pregnancies, but is further upregulated in preterm pregnancies complicated by chorioamnionitis, likely as a functional adaptation to protect the fetus from microbial and inflammatory cytokine entry into the intrauterine compartment by increasing placental efflux mechanisms. The placenta adapts its efflux mechanisms to protect the developing fetus from increased inflammation and infection present in preterm pregnancies^17,18^.

In our cohort, lower *IL-6* and unchanged *IL-8* mRNA expression in placentae of preterm pregnancies, including those with chorioamnionitis, may reflect a beneficial response to anti-inflammatory drug exposure to prevent PTB. *IL-6* plays a role in stimulating prostaglandin synthesis, an important hormone in the process of parturition, which may mediate the initiation of preterm labour^34^. In addition, IL-6 is involved in acute and chronic inflammation, assisting the immune response in the recruitment and activation of leukocytes^35, 36^, and involved in infection, where placental IL-6 mRNA^37^ and protein expression levels have been shown to increase with PTB and PTB associated with microbial invasion^38^. Previous studies have also shown elevated levels of IL-6 protein in the amniotic fluid^39^, maternal serum^37^ and placental decidual cells^40^ in pregnancies with chorioamnionitis. However, maternal medications administered during pregnancy in an effort to prevent PTB may reduce placental inflammation. Glucocorticoids can be used to reduce sterile intrauterine inflammation due to their anti-inflammatory properties, and umbilical cord plasma IL-6 concentrations have been shown to be lower in preterm pregnancies that received at least one dose of corticosteroids compared to those that received no glucocorticoids^41^. Tocolytics may be used to delay PTB and provide the opportunity to enhance fetal maturation through administration of corticosteroids, and tocolytics themselves also have anti-inflammatory properties^42, 43^. Tocolytics have been shown to suppress levels of pro-inflammatory mediators, including *IL-6* mRNA levels, in human placental cells stimulated by LPS and animal models of maternal infection^42^. Further, intrauterine infection may elicit an inflammatory innate immune response, which may contribute to preterm labour^44^, and antibiotics may reduce amniotic infection-associated inflammation^45^. In our cohort, mothers who delivered preterm and had chorioamnionitis received the greatest proportion of glucocorticoids, tocolytics, and antibiotics, followed by mothers who delivered preterm without chorioamnionitis, with term pregnancies receiving the lowest proportion of medications. Our data thus suggest that antenatal medications administered to prolong gestation and optimize offspring outcomes are successful in reducing some placental inflammation, though did not prevent preterm birth in this cohort.

Pregnancies with suboptimal maternal BMI may also be complicated by underlying inflammatory processes, however, in our cohort, maternal underweight and obesity did not affect the expression of placental MDR transporters or inflammatory cytokines. There was a small significant difference in placental mRNA expression levels of *ABCG2*, but not *ABCB1*, where placental mRNA expression of *ABCG2* augmented with increasing maternal BMI in preterm pregnancies. While there are a lack of data on the effect of maternal BMI on placental efflux transport function, animal studies have shown increased *Abcg2*, and decreased placental *Abcb1a* mRNA expression levels in high fat fed compared to undernourished pregnant mice^46^. In human pregnancy, maternal obesity has also been associated with downregulated placental *ABCB1* gene expression^47^. These alterations in placental efflux transporter expression in pregnancies complicated by maternal obesity could result in differences in placental transfer, with potential consequences for fetal drug exposure^48^. Maternal obesity has also been associated with increased levels of pro-inflammatory mediators in the placenta. In contrast to our findings, placental mRNA expression levels of *IL-6* have been shown to be increased in placentae from mothers with obesity compared to lean mothers^49, 50^. IL*-*8 has also been shown to be increased in placentae from women with obesity compared to non-obese placentae, but with no difference in *IL-6* placental expression^13^. Notably, our study criteria ensured the exclusion of maternal comorbidities such as gestational diabetes mellitus and hypertension, to better understand the effect of suboptimal BMI alone on outcomes. These comorbidities could be driving altered placental inflammation^51, 52^ and efflux transport^53, 54^ seen in other studies with maternal obesity and underweight. Including these conditions, and perhaps cases with more severe classes of maternal obesity, or a greater number of mothers who were underweight, may be needed to corroborate the effects of suboptimal maternal BMI on the placental pro-inflammatory phenotype.

Other strengths of our study include the cohort selection with corresponding clinical metadata; our study sample consists of data from 96 pregnancies, among term and preterm pregnancies (with and without chorioamnionitis), with further stratification by maternal underweight, normal weight, overweight, and obese BMI groups. This selection allowed for analysis not only by gestational age and infection status but by maternal BMI, which to our knowledge has not been previously done, with sufficient sample sizes. A limitation of our study is that only two select pro-inflammatory cytokines were measured, whereas the collective effect of multiple cytokines may be more indicative of the dynamic fluctuations in pro- and anti-inflammatory mediators during infection or labour. The cytokines selected for assessment have been previously demonstrated to be upregulated in PTB and maternal obesity^49, 50^, and may still be reflective of underlying inflammatory mechanisms in these conditions^40, 55^. Overall, our data will help to address gaps in knowledge on the impact of preterm birth, infection, and maternal prepregnancy BMI, in the absence of other maternal comorbidities, on placental-mediated protection of the fetus.

Our data show that preterm birth and infection status, but not maternal underweight or obesity, in otherwise uncomplicated pregnancies, alter placental expression of *IL-6* and MDR efflux transporters. Inflammation and infection have been associated with PTB and underweight/obesity, however, the underlying mechanisms linking these exposures to adverse pregnancy outcomes remain poorly understood. While current treatments for preterm delivery aim to delay the progression of contractions and eradicate infection, increased levels of inflammation *in utero* may still have consequences for fetal development. The upregulation of placental BCRP and P-gp at preterm may have implications for the biodisposition of drugs that could affect the developing fetus^33^, and should be considered in clinical treatment. Further, decreased placental *IL-6* may represent a beneficial response to anti-inflammatory medications administered for preterm labour. Altered placental expression of efflux transporters and inflammatory mediators may have implications for fetal exposure to xenobiotics in pregnancies complicated by PTB that could impair fetoplacental development.

## Data Availability

Data are available from the corresponding author upon reasonable request.

## Supplementary tables

**Supplementary Table 1.**
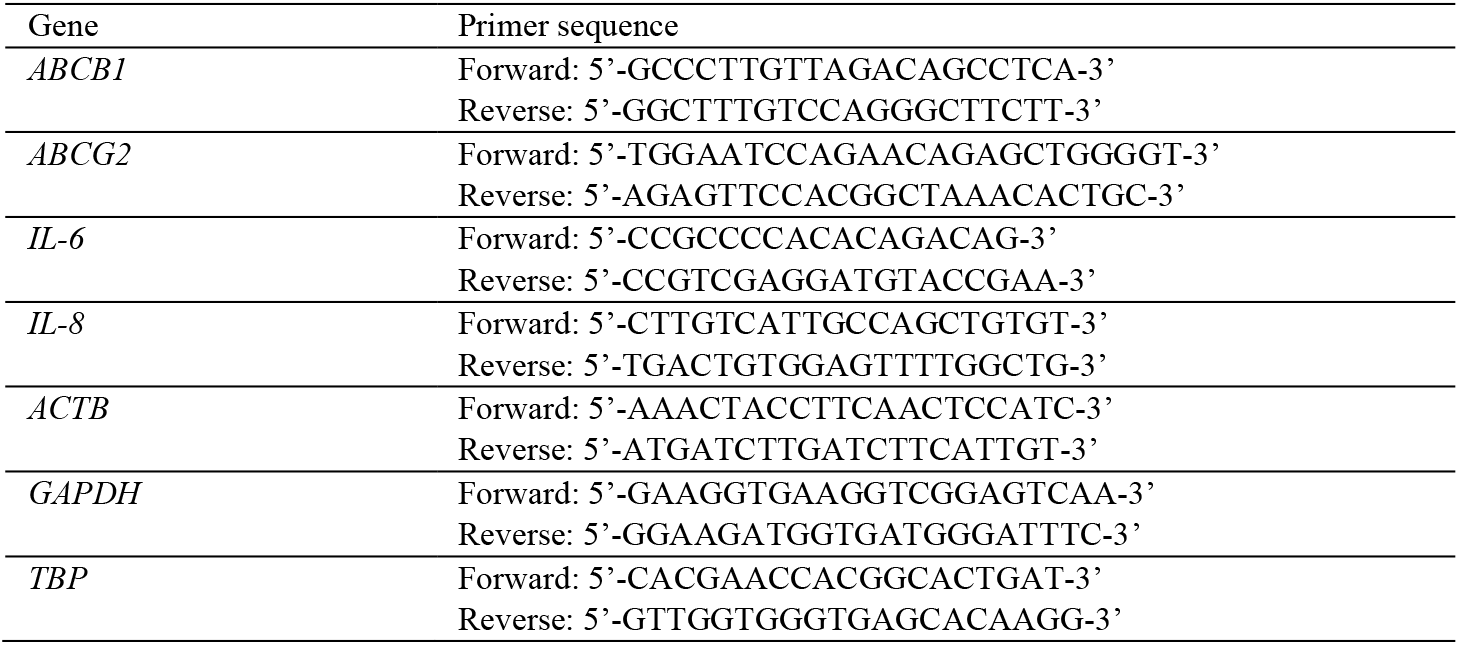
Primer sequences for real-time quantitative PCR gene studies.

**Supplementary Table 2.** Effect of maternal prepregnancy BMI on mRNA expression levels of placental MDR transporter and inflammatory genes in preterm and term pregnancies.

| Placental gene | Prepregnancy BMI |  |  |  | p-value |
| --- | --- | --- | --- | --- | --- |
|  | UW<br>(n=6-8) | NW<br>(n=6-16) | OW<br>(n=7-18) | OB<br>(n=8-11) |  |
| Preterm pregnancies |  |  |  |  |  |
| *ABCB1* | 1.14 (0.54, 1.32) | 1.10 (0.59, 1.28) | 0.91 (0.58, 1.21) | 0.66 (0.59, 1.05) | 0.64 |
| *ABCG2* | 0.62 ± 0.23 | 0.64 ± 0.19 | 0.67 ± 0.20 | 0.75 ± 0.19 | 0.45 |
| *IL-6* | 0.85 ± 0.29 | 0.82 ± 0.47 | 0.91 ± 0.36 | 0.96 ± 0.41 | 0.83 |
| *IL-8* | 0.18 (0.13, 0.84) | 0.33 (0.14, 1.09) | 0.55 (0.28, 0.84) | 0.22 (0.15, 0.44) | 0.23 |
| Term pregnancies |  |  |  |  |  |
| *ABCB1* | 0.65 (0.34, 1.33) | 0.67 (0.38, 1.01) | 0.61 (0.34, 0.76) | 0.75 (0.53, 1.81) | 0.30 |
| *ABCG2* | 0.76 ± 0.34 | 0.78 ± 0.23 | 0.70 ± 0.33 | 0.78 ± 0.29 | 0.95 |
| *IL-6* | 1.31 ± 0.50 | 1.24 ± 0.41 | 0.97 ± 0.19 | 1.38 ± 0.44 | 0.26 |
| *IL-8* | 0.26 (0.23, 0.48) | 0.22 (0.13, 0.62) | 0.25 (0.14, 0.34) | 0.64 (0.23, 0.82) | 0.21 |

Data are means ± SD (ANOVA; normal distribution/equal variance) or median (IQR; Kruskal-Wallis/Wilcoxon test for non-parametric data). \*p<0.05. UW = underweight. NW = normal weight. OW = overweight. OB = obese.

**Supplementary Table 3.** Effect of fetal sex on mRNA and protein expression levels of placental inflammatory genes and MDR transporters among preterm and term pregnancies.

|  | Preterm |  |  | Term |  |  |
| --- | --- | --- | --- | --- | --- | --- |
|  | Male<br>(n=34) | Female<br>(n=22) | p-value | Male<br>(n=14) | Female<br>(n=18) | p-value |
| Gene |  |  |  |  |  |  |
| *IL-6* | 0.93 ± 0.39 | 0.76 ± 0.32 | 0.14 | 1.25 ± 0.32 | 1.10 ± 0.30 | 0.22 |
| *IL-8* | 0.32 (0.15, 0.79) | 0.47 (0.18, 0.84) | 0.99 | 0.40 (0.22, 0.62) | 0.27 (0.16, 0.69) | 0.57 |
| Gene |  |  |  |  |  |  |
| *ABCB1* | 0.98 ± 0.47 | 0.92 ± 0.36 | 0.62 | 0.65 (0.39, 0.94) | 0.71 (0.38, 1.16) | 0.79 |
| *ABCG2* | 0.64 ± 0.19 | 0.72 ± 0.21 | 0.17 | 0.65 ± 0.23 | 0.83 ± 0.29 | 0.10 |
| Protein<sup>1</sup> |  |  |  |  |  |  |
| **P-gp** | 3 (1, 4) | 3 (1, 4) | 0.64 | 1.5 (0, 4) | 1 (0, 3) | 0.76 |
| **BCRP** | 4 (4, 4) | 4 (4, 4) | 0.75 | 4 (3.75, 4) | 4 (4, 4) | 0.72 |

Data are means ± SD (ANOVA; normal distribution/equal variance) or median (IQR; Kruskal-Wallis/Wilcoxon test for non-parametric data). \*p<0.05. <sup>1</sup>Immunolabeled area staining of the syncytiotrophoblast.

